# Evaluation of the causal effects of blood lipid levels on gout with summary level GWAS data: two-sample Mendelian randomization and mediation analysis

**DOI:** 10.1101/19006296

**Authors:** Xinghao Yu, Haimiao Chen, Shuiping Huang, Ping Zeng

**Author notes:** Correspondence to: Ping Zeng, Department of Epidemiology and Biostatistics, School of Public Health, Xuzhou Medical University, Xuzhou, Jiangsu, 221004, China; Center for Medical Statistics and Data Analysis, School of Public Health, Xuzhou Medical University, Xuzhou, Jiangsu, 221004, China, Email address.

## Abstract

**Objective:** Many observational studies have identified that gout patients are often comorbid with dyslipidemia, which is typically characterized by a decrease in high-density lipoprotein cholesterol (HDL) and an increase in triglycerides (TG). However, the relationship between dyslipidemia and gout is still unclear.

**Methods:** We first performed a two-sample Mendelian randomization (MR) to evaluate the causal effect of four lipid traits on gout and serum urate based on summary association statistics available from large scale genome-wide association studies (up to ∼100,000 for lipid, 69,374 for gout and 110,347 for serum urate). We adopted multivariable Mendelian randomization to estimate the causal effect independently. We also assessed the mediated effect by serum urate between lipids and gout with a mediation analysis. The MR results were validated with extensive sensitive analyses.

**Results:** Genetically lower HDL was positively associated with the risk of gout and serum urate concentration. Each standard deviation (SD) (∼12.26 mg/dL) increase was genetically associated with an odds ratio of gout of 0.75 (95% CI 0.62 ∼ 0.91, *p* = 3.31E-3) and with a 0.09 mg/dL (95% CI: -0.12 ∼ -0.05, *p* = 7.00E-04) decrease in serum urate concentration. Genetically higher TG was positively associated with the serum urate concentration. Each SD (∼112.33 mg/dL) increase was genetically associated with a 0.10 mg/dL (95% CI: 0.06 ∼ 0.14, *p* = 9.87E-05) increase in serum urate concentration. Those results were robust against various sensitive analyses. In addition, the multivariable Mendelian randomization confirmed the independent effect of HDL and TG on the gout/serum urate after adjustment for the other lipids. Finally, the mediation analysis showed that both HDL and TG could indirectly affect gout morbidity via the pathway of serum urate. The mediation effect accounted for about 13.0% or 28.0% of the total effect of HDL and TG, respectively.

**Conclusion:** Our study confirmed the causal associations between HDL/TG and gout/serum urate. Furthermore, the effect of HDL or TG on gout could also be mediated by serum urate.

**Key Messages:**

- Epidemiological studies have identified an accompanying association between lipid and gout. However, whether the association is causal is unclear.
- Mendelian randomization with genetic variants as instrumental variables is a useful tool facilitate the validation of a causal relationship for modifiable risk factors.
- The direct and indirect effects of lipids on gout, controlling for the serum urate concentration, can be estimated by a mediation analysis with serum urate serving as a mediator.
- We confirmed that elevated HDL levels can directly and indirectly lead to the decreased risk of gout, whereas elevation of TG levels can directly and indirectly elevate the risk of gout.

## Introduction

Gout is a common and complex form of arthritis with anyone at risk. It is characterized by sudden, severe attacks of pain, swelling, redness and tenderness in the joints, usually resulting from the chronic elevation of uric acid levels and the deposition of urate crystals in the body (e.g. joints and kidneys). Gout is manifested as rheumatism, gouty arthritis or gouty nephropathy, and is predominantly related to hyperuricemia due to metabolic disorders or reduced excretion of uric acid. Gout is typically an acute onset of single-joint synovitis in peripheral joints, causing extreme pain, whereas it is self-limiting and usually resolves within a few days (1-3). The global burden of gout is substantial, with the prevalence of gout ranging from 0.1% to 10% and the annual incidence from 30 to 600 cases per 100,000 individuals. In addition, gout is associated with several complex diseases (e.g. metabolic syndrome (4), cardiovasculopathy (5) and nephropathy (6)). Recently, gout per se was identified to be an independent risk factor in cardiovascular diseases (7, 8).

Beyond sustained hyperuricemia (9-11), many factors as genetic, environmental and social, e.g. diet, medication, comorbidities and exposure to heavy metals as cadmium and lead (12), are also involved in the pathogenesis of gout(13). In addition, some empirical studies have indicated a positive correlation between dyslipidemia and the risk of gout (14, 15), as evidenced in the increased prevalence and incidence of gout in people with dyslipidemia (16). Dyslipidemia is characteristic of aberrant blood lipids, including abnormalities of high-density lipoprotein cholesterol (HDL), low-density lipoprotein cholesterol (LDL), total cholesterol (TC) and triglycerides (TG). The third National Health and Nutrition Examination Survey of the United States reported that in gout patients the risk of hypertriglyceridemia was doubled and the odds ratio (OR) of low HDL was 1.60 after adjustment for age and gender (4), implying elevated HDL may lead to the decreased risk of gout. A cross-sectional study in 4,053 young adults revealed that the elevation of serum urate was associated with metabolic abnormalities (e.g. hyperinsulinemia, hypertension, dyslipidemia, and obesity) (17). In a study of the Japanese population, glucokinase regulatory protein was associated with decreased fasting blood glucose levels, elevated TG and serum urate concentrations, and was associated with dyslipidemia (18). On the contrary, some studies indicated that there was no significant difference in blood lipid levels between gout patients and normal controls (19).

These controversies over the relationship between dyslipidemia and gout may be partly attributable to uncontrolled/unmeasured confounders in observational studies. Additionally, doubt remains as to whether dyslipidemia is subsequent or consequent to gout. In case of limited observational evidence, Mendelian randomization with genetic variants as instrumental variables can facilitate the validation of a causal relationship for modifiable risk factors (20-22). In brief, Mendelian randomization holds that genetic variants associated with an exposure would also be associated with an outcome through the exposure route if the exposure is causally associated with the outcome.

Mendelian randomization employs genetic variants as instruments, and is often less susceptible to confounders compared with conventional observational studies in that genetic variants are randomly allocated at conception (23). This randomization process is analogous to the profile in a randomized controlled trial wherein participants are subjected to randomized classification and varied exposure. The substantial variation of outcome between groups provides evidence of the putative causal effect of the exposure on the outcome. In this sense, Mendelian randomization is at the interface of experimental and observational studies and is also referred to as the natural randomized trial (24, 25) to generate evidence in support of the potential causal effect of an exposure.

Indeed, Mendelian randomization has become a potent and effective statistical method for causal inference in observational studies, owing to recent achievements of genome-wide association studies (GWASs) (26-34). In the present study, we employed single nucleotide polymorphisms (SNPs) associated with lipid traits as instrumental variables to determine their causal associations with gout. We conducted the largest and most comprehensive two-sample Mendelian randomization analysis to date with summary statistics from large-scale GWASs with ∼180,000 individuals for lipid traits (i.e. HDL, LDL, TC and TG) (35), ∼2,100 cases for gout (36) and ∼110,000 individuals for serum urate (36).

## Materials and Methods

### GWAS data

The genetic data sets of the four lipid traits (i.e. HDL, LDL, TC and TG) were available from the Global Lipids Genetics consortium (GLGC) (35) (http://csg.sph.umich.edu/). After stringent quality control of genetic variants and individuals, ∼2,250,000 SNPs remained including genotyped and imputed from 188,577 individuals of European ancestry. Each of the four lipids was adjusted for available covariates (e.g. age, age^2^ and gender) and then the resultant residuals were quantile normalized to be a standard normal distribution. The association was finally performed for each SNP with a linear additive regression model. For each lipid trait we yielded its summary association statistics (e.g. effect allele, effect allele frequency, marginal effect size, standard error, p value and sample size).

We acquired genetic data sets for gout and serum urate concentration from the Global Urate Genetics Consortium (http://metabolomics.helmholtz-muenchen.de/) (36). After stringent quality control, a total of 69,374 (2,115 cases and 67,259 controls) European individuals and 2,538,056 genotyped and imputed SNPs remained for gout, and 110,347 individuals and 2,450,547 genotyped and imputed SNPs were reserved for serum urate. The association between each SNP and gout/serum urate was analyzed with an additive logistic/linear regression while adjusting for other available covariates (e.g. age and gender). Again, the summary association statistics of gout/serum urate (e.g. effect allele, marginal effect size, standard error, p value and sample size) were downloaded and refined. The GWAS genetic data sets used in our study are summarized in Table 1.

**Table 1.** Genetic data sets used in the present study

| Exposure(outcome) | Sample size(case/control) | Number of SNPs | Reference |
| --- | --- | --- | --- |
| HDL | 99,900 | 2,260,809 | (84) |
| LDL | 95,454 | 2,251,395 | (84) |
| TC | 100,184 | 2,259,768 | (84) |
| TG | 96,598 | 2,252,990 | (84) |
| Gout | 69,374 (2,115/67,259) | 2,538,056 | (36) |
| Serum urate | 110,347 | 2,450,547 | (36) |

HDL, High-density lipoprotein cholesterol; LDL, Low-density lipoprotein cholesterol; TC, Total cholesterol; TG, Triglycerides.

### Selection of instrumental variables

Based on our prior studies (28, 37), we selected instrumental variables for lipids with the use of the clumping procedure of PLINK (version v1.90 b3.38) (38). When clumping, we set both the primary significance level and the secondary significance level for index SNPs to 5.00E-8, the linkage disequilibrium and the physical distance to 0.01 and 1,000 kb, respectively. We then selected the lipid-specific index SNPs as instrument variables and finally generated 103 SNPs for HDL, 50 SNPs for LDL, 60 SNPs for TC and 60 SNPs for TG. As per previous MR literature (28, 31, 39-48), we further examined pleiotropic association by removal of SNPs which may be potentially associated with gout or serum urate. Specifically, we excluded index SNPs which had an adjusted p value of less than 0.05 after Bonferroni correction or residing within the gout- or serum-urate-associated loci (i.e. 1 Mb in the physical distance). Of note, the removal of SNPs with pleiotropic effects is an authenticated manner to ensure the validity of Mendelian randomization (37).

### Estimation of causal effect and sensitivity analyses

We subsequently employed Mendelian randomization to determine the causal relationship between lipids and gout/serum urate. First, we calculated the proportion of phenotypic variance explained (PVE) by instruments and computed the *F* statistic for the four blood lipids to quantitatively verify whether the selected index SNPs were strong instruments (28, 37, 49, 50). For the four lipid traits, we initially conducted the inverse variance weighted (IVW) method to estimate their causal effects on gout/serum urate (51). We then conducted several sensitivity analyses to assess the validity of our IVW Mendelian randomization results: (1) weighted median-based method (52) and maximum likelihood method (53); (2) leave-one-out (LOO) cross-validation analysis (37, 49) to validate whether there were instrumental outliers that can substantially impact the causal effect estimates; (3) MR-Egger regression to further evaluate horizontal pleiotropic effects of instrumental variables (54); (4) reverse causal analysis to assess whether gout/serum urate exhibited a causal effect on lipids using instrumental variables of gout or serum urate, since the determination of the causal direction is very important in Mendelian randomization (55); (5) multivariable Mendelian randomization analysis (28, 37, 56) to investigate the relationship between one lipid trait (e.g. HDL) and gout/serum urate while adjusting for the effects of other lipids (e.g. LDL, TC and TG).

### Mediation analysis to explore the mediation effect of serum urate in the path from lipids to gout

Emerging evidence demonstrated that elevated serum urate is the most important single-risk factor and urate-lowering therapy is frequently recommended in the clinical management of gout patients (57). Nonetheless, on the grounds of the causal relationships between lipids and serum urate identified in our analysis, a natural and immediate problem arose as to whether serum urate can mediate the effect of lipids on gout (Fig. 1 C), or rather, do lipids have an indirect effect on gout via serum urate? To address this problem, we further performed a mediation analysis with serum urate serving as a mediator and estimated the mediating effect of serum urate (i.e. the indirect effect of lipids) (58-62). The estimation and hypothesis testing of the mediation effect were also implemented within the framework of Mendelian randomization based on summary association statistics of lipids, serum urate and gout (Table 1 and Fig. 1 D). Briefly, we estimated the causal effect of lipids on serum urate with IVW methods, and estimated the causal effect of serum urate on gout by the multivariable Mendelian randomization analysis. Such an analysis is also referred to as network Mendelian randomization (63, 64). The mediation effect was evaluated with the product method and was tested with the Sobel test (65) or the Bootstrap test (66). The details of the mediation analysis were demonstrated in Additional file 1.

Our statistical analysis was mainly conducted within the R (version 3.5.2) software. As there were four exposures and two outcomes in our Mendelian randomization analysis, the statistical significance level was adjusted to 6.30E-03 (= 0.05/8) to incorporate the issue of multiple hypothesis testing. In addition, since participants had provided written informed consent for data sharing as described in each of the original GWASs; ethical review was omitted in our study.

## Result

### Causal effect of lipids on gout

We screened out 92, 48, 58 and 53 lipid-specific instrumental variables for HDL, LDL, TC and TG when evaluating their associations with gout. Together, these instrumental variables accounted for a total of 5.7%, 3.1%, 2.6 % and 4.3% phenotypic variances for HDL, LDL, TC and TG, respectively. The *F* statistics for all these SNPs were greater than 10 (Additional file 2: Tables S1-S4), suggesting that weak instrument bias was potentially null in our analysis. The Cochran’s Q test (67, 68) showed that there was little evidence of instrumental heterogeneity for HDL (*p* = 0.945), TC (*p* = 0.179) or TG (*p* = 0.379). However, an instrumental heterogeneity was observed for LDL at the marginal significance level of 0.05 (*p* = 0.047). Therefore, we employed the fixed-effects IVW method for HDL, TC and TG, and applied the random-effect IVW method for LDL when estimating the causal effects on gout.

Among the four lipids we identified that only HDL was associated with gout (Fig. 2A-C and Fig. 3). Specifically, the estimated odds ratio (OR) per standard deviation (SD) increase of HDL (∼12.26 mg/dL) on gout was 0.75 (95% CI 0.62 ∼ 0.91, *p* = 3.31E-3) and was statistically significant after Bonferroni correction. The OR per SD increase of LDL (∼30.25 mg/dL), TC (∼36.32 mg/dL) or TG (∼112.33 mg/dL) for gout was 0.81 (95% CI 0.61 ∼ 1.07, *p* = 0.079), 0.98 (95% CI 0.90 ∼ 1.06, *p* = 0.604) and 1.16 (95% CI 0.93 ∼ 1.45, *p* = 0.186) (Fig. 3 and Additional file 3: Table S5), respectively.

### Causal effect of lipids on serum urate

We generated 87, 45, 57 and 49 lipid-specific instrumental variables for HDL, LDL, TC and TG when evaluating their associations with serum urate. These instrumental variables explicated a total of 5.0%, 3.0%, 2.5 % and 3.7% phenotypic variances for HDL, LDL, TC and TG, respectively (Additional file 2: Tables S6-S9). Thereafter, we used the random-effects IVW method to estimate the causal effects due to the heterogeneity observed for all the four sets of instruments (the *p* values of the Cochran’s Q test were 5.37E-10, 6.95E-04, 3.41E-06 and 2.67E-03 for HDL, LDL, TC and TG, respectively). HDL is negatively associated with serum urate after Bonferroni correction (estimated causal effect = -0.09, 95% CI -0.12 ∼ -0.05, *p* = 7.00E-04) (Fig. 2D-F and Fig. 3). In addition, a positive association existed between TG and serum urate (estimated causal effect = 0.10, 95% CI 0.06 ∼ 0.14, *p* = 9.87E-05) (Fig. 2G-I and Fig. 3). However, no association was observed between LDL/TC and serum urate (Additional file 3: Table S5 and Fig. 3).

### Sensitivity analyses to validate the estimated causal effects

We subsequently performed sensitivity analyses to validate the causal association observed above. Herein, we relegated the results of serum urate to Additional file 4 and focused only on gout in the following paragraphs. Due to the insignificant association between LDL/TC and gout/serum urate, we only summarized their results in Fig. 3 and Additional file 3: Figures. S1-S3, but did not pursue any of these two sets of traits further.

With the maximum likelihood method, the estimated OR per SD increase of HDL on gout was 0.75 (95% CI: 0.62 ∼ 0.91, *p* = 3.31E-03), in line with the IVW estimate; while the weighted median method identified a consistently inverse but insignificant association (OR = 0.81, 95% CI 0.59 ∼ 1.11, *p* = 0.197). The LOO analysis showed that none of the instruments of HDL could individually affect the substantial causal effect estimates (Additional file 3: Figure S4). However, Fig. 2A delineated that two instrumental variables of HDL with large effect sizes on gout seemed to be potential outliers (i.e. rs2566091 in gene *ALDH1A2* and rs9989419 in gene *AC012181*.*1*). However, they did not exert any substantial impact on the estimated causal effect of HDL on gout. Specifically, after removal, the OR was 0.74 (95% CI 0.60 ∼ 0.91, *p* = 4.18E-03), similar to that obtained with the instruments in total (Fig. 2B). Furthermore, the funnel plot of HDL for gout exhibited a symmetric pattern around the causal effect point estimates (Fig. 2C), offering little evidence for horizontal pleiotropy. The MR-Egger regression also precluded the possibility of pleiotropy of instrument variables (the intercept = -0.013, 95% CI -0.028 ∼ 0.003, *p* = 0.113). We further performed a multivariable Mendelian randomization analysis to estimate the association between HDL and gout while adjusting for LDL, TC and TG. The result showed HDL a de facto inverse relationship with gout even after controlling for the residual three lipid traits (Additional file 3: Table S10), implying the independent causal role of HDL in the risk of gout.

In the reverse causal analysis, we kept one instrumental variable for gout in estimation of its causal effect on HDL, and 24 (or 22) instrumental variable of serum urate when estimating its effect on HDL (or TG) (Additional file 2: Tables S11-S13). With the fixed-effects IVW method, the causal effect of gout on HDL was estimated to be -0.019 (95% CI: -0.018 ∼ 3.20E-03, *p* = 0.173), and the causal effects of serum urate on HDL and TG were estimated to be -0.033 (95% CI -0.065 ∼ -7.31E-04, *p* = 0.045) and 0.016 (95% CI: -0.016 ∼ 0.048, *p* = 0.330), respectively. All of estimates were insignificant in case of multiple hypothesis testing. Therefore, this analysis excluded the probability of reverse causation from gout/serum urate to lipids, indicating that dyslipidemia (e.g. decreased HDL and/or increased TG) was a causal factor rather than a clinical manifestation of gout or serum urate.

### Results of the mediation analysis

To estimate the possible mediation of serum urate in the progression from dyslipidemia and gout, we combined both the instrumental variables of lipids and serum urate, and reserved 116, 65, 76 and 73 lipid-specific instrumental variables for HDL, LDL, TC and TG in the mediation analysis (Additional file 2: Tables S14-S18). Notably, the following reported effects were corrected for the same scales (Additional file 1).

In the mediation analysis we observed that the summation of the mediation effect and the direct effect was approximately equal to the total effect (Table 2), indicating that our correction strategy used for those effects was justifiable. The total effect of HDL on gout was estimated to be -0.154 (95% CI: -0.256 ∼ -0.051, *p* = 3.31E-3; Fig. 3) and the direct effect of HDL on gout was estimated to be -0.106 (95% CI: -0.189 ∼ -0.022, *p* = 1.35E-02). More importantly, the mediation effect of HDL on gout was estimated at -0.020 (95% CI: -0.033 ∼ -0.008, *p* = 1.67E-03), which accounted for about 13.0% (= 0.020/0.154) of the total effect, indicating that serum urate was a promising mediator between HDL and gout, and that HDL could indirectly affect the risk of gout via serum urate in addition to the direct impact. Likewise, the total effect of TG on gout was estimated at 0.082 (95% CI: -0.039 ∼ 0.202, *p* = 1.86E-01) and the direct effect of TG on gout was estimated at 0.048 (95% CI: -0.057 ∼ 0.152, *p* = 3.71E-01). Furthermore, the mediation effect of TG on gout was estimated at 0.023 (95% CI: 0.010 ∼ 0.037, *p* = 8.39E-04), accounting for about 28.0% (= 0.023/0.082) of the total effect. TG also served as a mediator between TG and gout and could indirectly increase the risk of gout via serum urate. However, the mediation effects of serum urate between LDL/TC and gout were insignificant (Table 2).

**Table 2.**
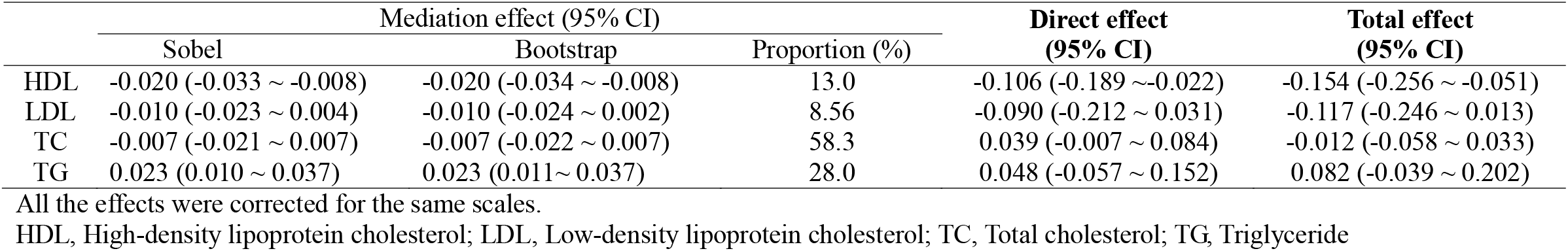
Mediation analysis of lipids on gout with serum urate concentrations

## Discussion

### A summary of our study

Many observational studies have reported that gout patients are often comorbid with dyslipidemia (15, 69-72). However, the relationship between the two clinical disorders still remains masked. To explore whether the association between dyslipidemia and gout is causative, we performed a two-sample Mendelian randomization analysis in the present study. To our best knowledge, this is the first study that attempts to investigate the causal relationship between lipid/serum urate and gout by employment of summary association statistics obtained from large-scale GWASs with Mendelian randomization within the framework of instrumental variable approaches (20).

With the IVW method, we observed that HDL was negatively associated with gout and TG positively associated with serum urate. These inferred causal relationships were robust against the designation of statistical methods. A series of sensitivity analyses (e.g., MR-Egger regression) excluded the probability of instrumental pleiotropy that can contribute to bias in the causal effect estimation. We also removed the reverse causality and confirmed that the variations of HDL and TG levels were the premises of gout/serum urate rather than consequences. Furthermore, the multivariable Mendelian randomization confirmed the independent effect of HDL and TG on the gout/serum urate after adjustment for the other lipids. With a mediation method, we identified a mediation effect between HDL and gout mediated by serum urate, which indicated that elevated HDL could decrease the serum urate and, in addition to the direct influence, indirectly relieved the risk of gout. Moreover, a positive mediation effect was detected between TG and gout, i.e., though not a causal contributor of gout, TG could indirectly exacerbate the risk of gout by elevating serum urate. Additionally, we confirmed the positive association of serum urate with gout, which indicated that elevated serum urate concentrations can causally increase the risk of gout. Overall, our study affirmed that elevated HDL levels can directly and indirectly lead to the decreased risk of gout, whereas elevation of TG levels can directly and indirectly elevate the risk of gout.

### Mechanism underlying the association between dyslipidemia and gout/serum urate

The association between dyslipidemia and gout/serum urate is very complicated, implicated in a number of mechanisms. Dyslipidemia may lead to an increase in ketone body and reduce the acid-discharging ability of the kidneys, with the renal arteries and the micro-arteries of the glomeruli impaired. Consequently, stenosis and occlusion of the diseased blood vessels develop, with the excretion of uric acid from the kidneys is largely discounted (73). Furthermore, hyperuricemia is reportedly associated with metabolic syndrome, which is characteristic of hyperinsulinemia due to insulin resistance. An increase in TG or a decrease in HDL may result in insulin resistance, consequently leading to the increased serum urate during glycolysis and free fatty acid metabolism (74). Meanwhile, due to the increased reabsorption of uric acid by the kidneys, serum urate directly contribute to hyperuricemia (75).

The negative association between HDL and gout/serum urate may be attributable to the anti-atherosclerotic, anti-oxidative, anti-inflammatory and anti-thrombotic effects of HDL (76-78). Our mediation analysis revealed that, beyond the direct effect, HDL can directly abate the risk of gout by decreasing serum urate concentration. The positive correlation between TG and serum urate may be exemplified by the finding that TG promotes the synthesis whereas decreases the normal excretion of serum urate. Indeed, TG can indirectly affect the gout via serum urate. Since excessive intake of TG-rich food (e.g. fructose and fat-rich food) may cause hyperthyroidism and further increase uric acid levels (79), overmuch fat metabolism-related products will inhibit the excretion of serum urate. Furthermore, the synthesis of triglycerides requires NADPH, which possibly increases the concentration of serum urate (80). Intriguingly, clinical studies have validated that patients with hyperuricemia or gout have a 19% reduction in serum urate concentration at end of a three-week regimen of oral lipid-lowering drugs, and serum urate concentration rebounded to its original level after discontinuation of the agents (81), the mechanism for which awaits further investigation.

### Limitations

Like other Mendelian randomization methods, our research has certain limitations. First, as mentioned above, due to the small sample size of cases in gout GWAS, the statistical power in our Mendelian randomized analysis was limited. For example, in our study, the sample size of adult gout was 70,000 and the proportion of cases was only 3.1%; the power calculation results showed that we had a small to moderate power to detect the causal association between lipids and gout (Fig. 4)(82). The estimated statistical power was 28% or 15% to detect an OR of 0.70/1.30 or 0.80/1.20, respectively. Second, we hypothesized that the relationship between blood lipid levels and gout was linear in our analysis, but this linear association might not be present in the clinical scenario, and thus we could not completely eliminate the non-linear effects of lipids on gout. Third, as individual-level datasets were not available; therefore, we could not evaluate the effects of extreme lipid levels on gout/serum urate or conduct a stratified analysis (e.g. gender). In addition, again due to the unavailability of individual-level data, we could not investigate the relationship between the TG-HDL ratio, a known indicator of insulin resistance (83), and gout. Fourth, we adopted the multivariable Mendelian randomization method to remove the effects of pleiotropic effects, but this method was not applicable to the unknown or unmeasured pleiotropy (56).

## Conclusion

In conclusion, our study confirmed the causal associations between HDL/TG and gout/serum urate levels. Furthermore, serum urate can serve as a mediator for the effect of HDL or TG on gout.

## Data Availability

We are indebted to the GLGC and Global Urate Genetics Consortium studies for public availability in making the summary data and we are grateful to all the investigators and participants for their contributions to those studies.

## Abbreviations

HDL: High-density lipoprotein cholesterol
GLGC: Global Lipids Genetics consortium
GWASs: genome-wide association studies
IVW: inverse variance weighted
LDL: Low-density lipoprotein cholesterol
LOO: leave-one-out
MR: Mendelian randomization
OR: odds ratio
PVE: phenotypic variance explained
SD: standard deviation
SNPs: single nucleotide polymorphisms
TC: Total cholesterol
TG: Triglyceride.

## Acknowledgements

We are indebted to the GLGC and Global Urate Genetics Consortium studies for public availability in making the summary data and we are grateful to all the investigators and participants for their contributions to those studies. The data analyses in the present study were supported by the high-performance computing at Xuzhou Medical University.

## Authors’ Contributions

PZ and SH conceived the design of the study; PZ and XY obtained the data; PZ and XY cleared up the datasets; PZ and XY mainly performed the data analyses; HC, YG and JY helped clear and analyze the data; PZ, XY and FG interpreted the results of the data analyses; PZ and XY drafted the manuscript, and all authors approved the manuscript and provided relevant suggestions.

## Disclosure

The authors declare that the research was conducted in the absence of any commercial or financial relationships that could be construed as a potential conflict of interest.

## Funding

This work was supported by Youth Foundation of Humanity and Social Science funded by Ministry of Education of China (18YJC910002), the Natural Science Foundation of Jiangsu (BK20181472), the General China Postdoctoral Science Foundation (2018M630607), the Special China Postdoctoral Science Foundation (2019T120465), the Postdoctoral Science Foundation of Xuzhou Medical University, QingLan Research Project of Jiangsu for Outstanding Young Teachers and Six-Talent Peaks Project in Jiangsu Province of China (WSN-087), the National Natural Science Foundation of China (81402765), the Statistical Science Research Project from National Bureau of Statistics of China (2014LY112), the Postgraduate Research & Practice Innovation Program of Jiangsu Province (KYCX19_2250), Social Development Project of Xuzhou, and the Priority Academic Program Development of Jiangsu Higher Education Institutions (PAPD) for Xuzhou Medical University.

## Additional file

**Additional file 1**: Supplementary Methods;

**Additional file 2:** Supplement Tables;

**Additional file 3:** Supplement Tables and Figures;

**Additional file 4:** Supplementary Results.

## Notes

### Competing Interest Statement

The authors have declared no competing interest.

### Author Declarations

All relevant ethical guidelines have been followed and any necessary IRB and/or ethics committee approvals have been obtained.

Any clinical trials involved have been registered with an ICMJE-approved registry such as ClinicalTrials.gov and the trial ID is included in the manuscript.

